# Clinical phenotype of familial hypertensive nephropathy

**DOI:** 10.64898/2026.06.23.26356313

**Authors:** Guy H. Neild, D. Deren Oygar, Ahmet Behlul, Salahi Ataç, Meral Yükseliş, Simge Özadalı, Fezile Ozdemir, Hasan Hüseyin Kazan, Daniel P. Gale, Cemal Gurkan

**Author notes:** **Corresponding Author:** Professor Guy Neild, University College London Centre for Kidney and Bladder Health, Royal Free Hospital, Rowland Hill Street, London, NW3 2PF, UK.

## Abstract

Familial kidney disease is common in Cyprus. Patients with a glomerular phenotype are most likely to have an autosomal dominant variant in collagen type IV alpha 3 chain (*COL4A3*) or collagen type IV alpha 4 chain (*COL4A4*) genes but pathogenic variants are not found in the majority of families. We compare the clinical phenotype between two groups of 10 Turkish Cypriot families who lack a pathogenic variant of *COL4A3* or *COL4A4* but have either the *COL4A4* variant p.G545A or p.G999E. Both groups had identical clinical phenotypes with microscopic haematuria detected at least once in 76% of affected family members; urine protein was less than 1 g/day until glomerular filtration rate (GFR) was <30 ml/min. End-stage kidney disease (ESKD) occurred in 24.1% of those over 50 at a median age of 62.8 (36-86) years. Although the genetic cause of renal injury in this large group is still unknown, these families present with a clinical phenotype best characterised as familial hypertensive nephropathy. We propose that this condition accounts for the great excess of renal failure in the Eastern Mediterranean in those over 65 years of age.

## Introduction

Chronic kidney disease (CKD) is common in Cyprus and the Eastern Mediterranean [1], and the incidence of renal failure is 3-times more common than that in the Northern Europe [2]. Familial kidney disease is also common and earlier investigation of Greek Cypriot families by Deltas and his colleagues has shown that the majority of families with CKD have a mild glomerular phenotype with a haematuric nephropathy usually as a consequence of autosomal dominant (AD) variants in COL4A3 and COL4A4 [3–5]. The nephropathy is characterized in adolescence and early adult life by microscopic haematuria and electron microscopy of renal biopsy specimens show thin glomerular basement membranes (GBM). Subsequently, over the age of 30 a significant percentage develop CKD and end-stage kidney disease (ESKD). Proteinuria with CKD developed in 8% of all mutation carriers between 31 and 50 years. Of those with proteinuria, 74% subsequently developed CKD and of these, 43% reached ESKD at a mean age of 60 years. Histology of those with progressive renal failure showed advanced segmental glomerulosclerosis [3,4].

The *COL4A3*, *COL4A4* and *COL4A5* genes code for the collagen IV alpha 3-5 chains that form the major collagen IV alpha 3, alpha 4 and alpha 5 network of the basement membranes in the kidney, ear and eye [6]. Although the X-linked Alport syndrome and autosomal recessive (AR) forms are well recognized until recently AD form was thought to be confined to a syndrome of benign familial haematuria and with thin glomerular basement membrane nephropathy (GBM) as measured by electron microscopy (EM) [7].

Our own investigation of Turkish Cypriots has confirmed that pathogenic variants in *COL4A3* or *COL4A4* can be found in families with a conspicuous glomerular phenotype but in the majority of families no pathogenic variants are found [5,8]. On review of probands without any probable pathogenic variant, we noted that two *COL4A4* variants, NM_000092.5: c.1634G>C:p.Gly545Ala (p.G545A) and NM_000092.5: c.2996G>A:p.Gly999Glu (p.G999E), with likely-pathogenic scores occurred frequently. Both the p.G545A and p.G999E variants are common with allele frequencies in gnomAD v4.1 of 0.0817 and 0.012 respectively and are classified as benign by American College of Medical Genetics and Genomics (ACMG) but nevertheless show several characteristics of a pathogenic variant. For instance, both are SIFT 0, PolyPhen 1.0, Revel 0.776 and 0.773, Grantham scores 60 and 98 respectively. ACMG *in silico* predicts an aggregated score of 0.837 with a deleterious effect [8]. The role or relevance of these two variants is unknown as (i) families with CKD without these variants appear no different, (ii) the variants do not always co-segregate with the affected members and (iii) those with homozygous copies do not appear to be at any disadvantage.

We have previously reported the clinical phenotype of five families with the p.G545A variant, which we described as similar to ‘hypertensive nephropathy’ and more typical of a tubular disease until patients reach CKD stage 4 (eGFR <30 ml/min) [8]. Now we report the clinical phenotype from 20 families with no pathogenic variant, in whom the renal condition shows an autosomal dominant pattern. For convenience, we compare the clinical phenotype in two groups of 10 families with either the p.G545A or p.G999E variant. We expect the two variant groups to be similar and more typical of a tubular disease until CKD stage 4 is reached.

## Materials and Methods

This is a retrospective study of the natural history of progressive kidney disease in Turkish Cypriot families. Hospital records were reviewed during the period 1 January 2024 to 31 May 2026.

Written informed consent was obtained for participation in this study. All sample collections were performed with written informed consent and in accordance with the ethical principles for medical research involving human subjects and according to the Declaration of Helsinki of the World Medical Association. Patients and family members donated DNA between March 1^st^ 2019 and 31^st^ December 2025.

### Families

The study was conducted at Dr. Burhan Nalbantoglu Nicosia State Hospital in Northern Cyprus, a tertiary care hospital that provides all renal services to Northern Cyprus. Family trees were created for families with 2 or more members with evidence of chronic kidney disease (CKD) [9].

Families were contacted and were either visited in their villages or seen at the hospital. Family trees have been accumulated since 2009 over a 17-year period. The majority of families have an AD inheritance. Our work has been helped by the number of large families with 6–10 siblings in early generations.

After providing written informed consent, all volunteer family members underwent urinalysis and renal function evaluation, and DNA was donated.

### DNA investigations

DNA extractions were carried out using the PureLink Genomic DNA Mini Kit (Invitrogen, Carlsbad, CA, USA) according to the manufacturer’s procedure. All DNA samples were quantified using a Qubit 4.0 fluorometer and a Qubit dsDNA HS Assay Kit (Invitrogen, Carlsbad, CA, USA).

*COL4A3* and/or *COL4A4* variants were sought either by whole exome sequencing (WES), or next generation sequencing (NGS) with parallel analysis of a 5-gene panel containing *COL4A3*, *COL4A4*, *COL4A5*, complement factor H related 5 (*CFHR5*), and fibronectin 1 (*FN1*) as previously described [8].

Families with CN had at least one member examined by real-time polymerase chain reaction (RT-PCR) for the two variants p.G545A and p.G999E [8].

### Study groups investigated

A total of 118 families has been investigated, of whom 72 probands underwent WES, 57 underwent NGS and 11 had both investigations.

To describe the phenotype of CN, we have summarized the clinical information and analyzed 10 families with p.G545A, and 10 families with the p.G999E variant, in whom there were at least 6 clinically affected members.

### Clinical investigations

Kidney function (eGFR) was reported by the Modification of Diet in Renal Disease (MDRD) formula. The upper limit of normal for creatinine in men was 97 µmol/l, for women 80 µmol/l, and for blood urea nitrogen (BUN) 7.1 mmol/l.

Proteinuria was initially measured by 24-hr urine collection, and a normal protein concentration defined as <140 mg/day and an albumin concentration <30 mg/day. More recently, proteinuria has been measured as the protein and albumin/creatinine ratio (urine albumin-to-creatinine ratio (uACR) <30 mg/mg and urine protein-creatinine ratio (uPCR) <200 mg/mg).

Haematuria was investigated historically by urine microscopy and more recently by urine dipstick tests. Haematuria was deemed positive and equivalent to ‘a trace’ on a dipstick if there were 3-4 red blood cells per high-power field.

Eleven kidney biopsies have been performed in 10 families. Biopsy specimens were routinely examined by light microscopy and immunofluorescence, and in one case electron microscopy data were available.

A modified CKD staging was used: stage 0, no evidence of kidney disease; stage 1, eGFR>90 and microscopic hematuria; stage 2, eGFR 60-89 ml/min, microscopic hematuria and/or uACR >30 mg/mg; stage 3, eGFR in the range of 59-30 ml/min; stage 4, eGFR 29-15; and stage 5, kidney failure with eGFR <15 ml/min.. The diagnostic criteria were based on Kidney Disease: Improving Global Outcomes (KDIGO) guidelines [9].

### Statistical analysis

Age at onset of CKD and ESKD, and percentage reaching end-stage after age 50 were compared by t-test.

## Results

### Families with no pathogenic variant

From a total of 118 families, we have identified 10 families with p.G545A and 10 with p.G999E, and with at least six affected members in two or more generations.

In the 20 families reviewed, 581 members were recorded, 215 have evidence of CKD (Stages 1-5), 131 have reached Stage 3 (eGFR <60 ml/min), and 45 have reached ESKD (Stage 5; Table 1).

**Table 1:**
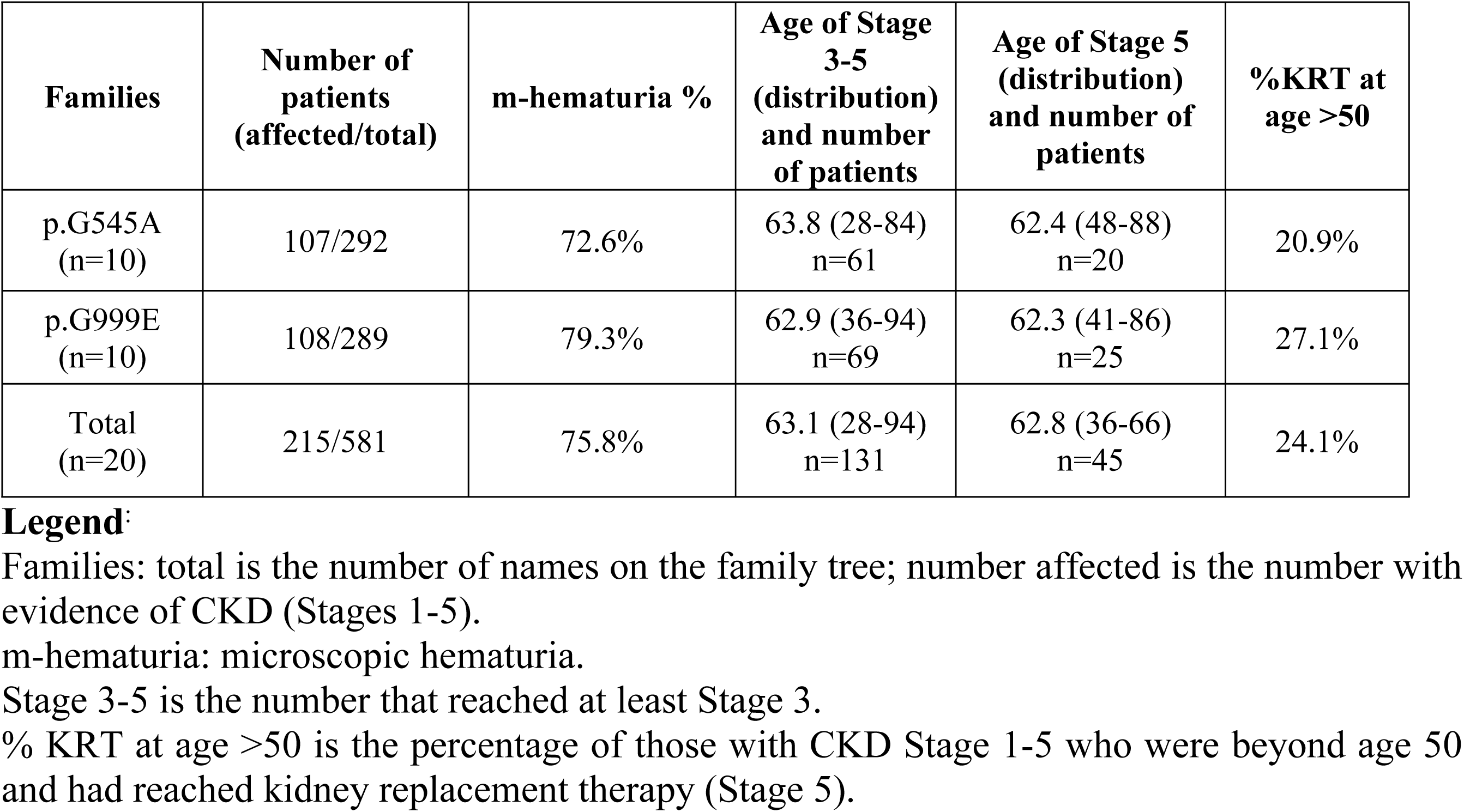
Demographic details of the patients.

| Families | Number of patients (affected/total) | m-hematuria % | Age of Stage 3-5 (distribution) and number of patients | Age of Stage 5 (distribution) and number of patients | %KRT at age >50 |
| --- | --- | --- | --- | --- | --- |
| p.G545A (n=10) | 107/292 | 72.6% | 63.8 (28-84) n=61 | 62.4 (48-88) n=20 | 20.9% |
| p.G999E (n=10) | 108/289 | 79.3% | 62.9 (36-94) n=69 | 62.3 (41-86) n=25 | 27.1% |
| Total (n=20) | 215/581 | 75.8% | 63.1 (28-94) n=131 | 62.8 (36-66) n=45 | 24.1% |

Families: total is the number of names on the family tree; number affected is the number with evidence of CKD (Stages 1-5).
m-hematuria: microscopic hematuria.
Stage 3-5 is the number that reached at least Stage 3.
% KRT at age >50 is the percentage of those with CKD Stage 1-5 who were beyond age 50 and had reached kidney replacement therapy (Stage 5).

Of the affected members: mildly raised blood pressure was universal by CKD stage 3 and was easily controlled with 1-2 drugs. There was no case of accelerated hypertension considered relevant to deteriorating kidney function.

Microscopic hematuria could be intermittent or absent. Among the 117 people whose urine was tested on one or more occasions, 89 had a positive result at least once (76%).

We found that the dipstick test for proteinuria was 0 to 1+ until the eGFR was less than 30 ml/min. Proteinuria versus eGFR is shown in Fig 1, documenting the observation that proteinuria was not greater than 1 g/d until the eGFR was less than 30 ml/min. We noted that for proteinuria (more than 140 mg/d) in patients with an eGFR >50 ml/min, approximately 50% of the urine protein was albumin, and this percentage increased to 80-90% when the proteinuria exceeded 2.0 g/d with eGFR values less than 30 ml/min.

**Figure 1.**
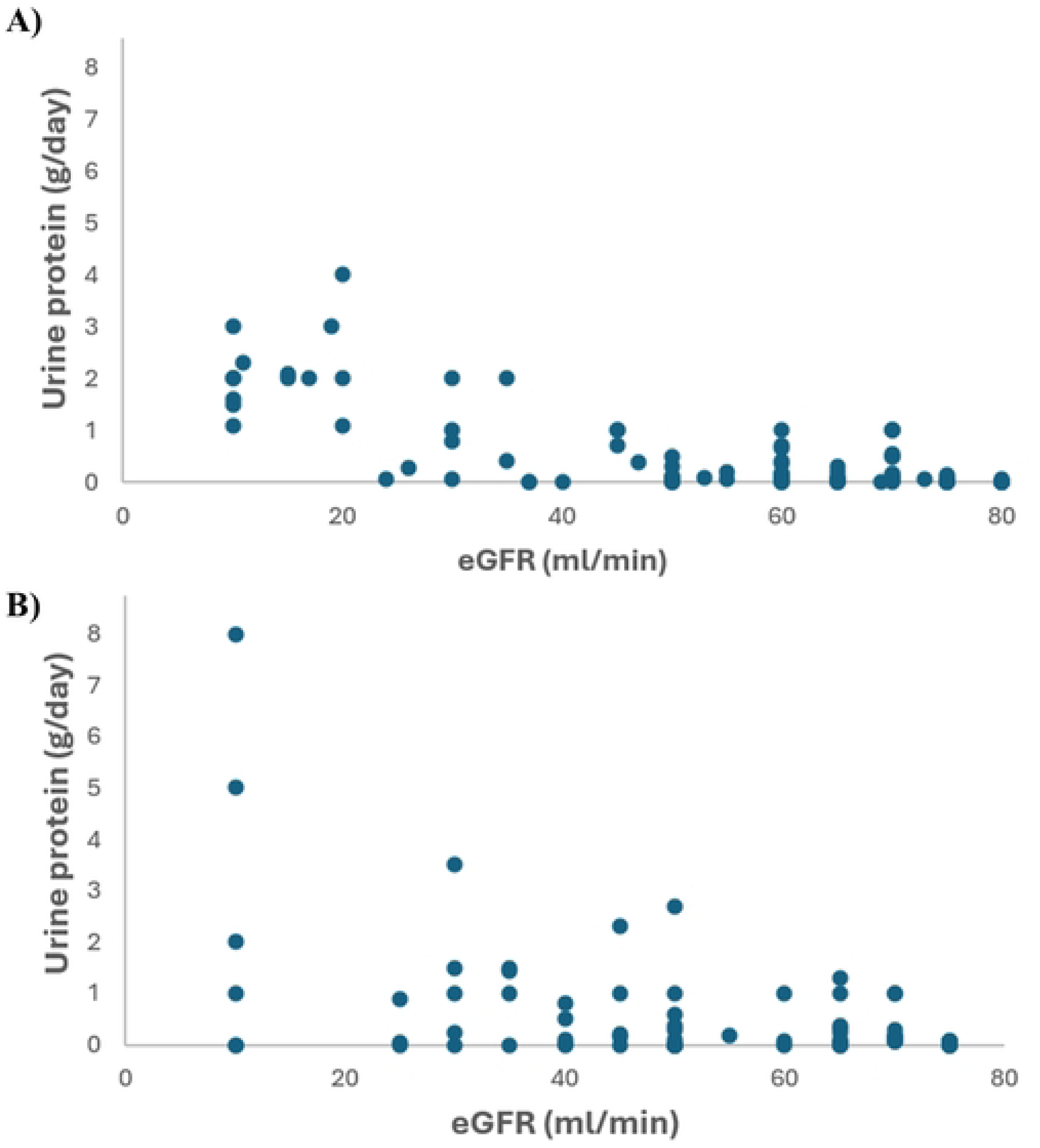
Proteinuria vs eGFR levels for patients with A) p.G545A and B) p.G999E variants.

The age of onset of CKD stage 3 and ESKD is shown in Table 1. Among family members with clinical evidence of renal disease (stage 1-5) and aged older than 50 years, 24.1% had progressed to ESKD by a median age of 62.8 years (range 36-86 years).

### Biopsies

Eleven biopsies were performed in 10 families without a pathogenic variant. All biopsy specimens were described as either secondary focal and segmental glomerulosclerosis (2° FSGS) or hypertensive nephropathy. Reports on six are available and show the characteristic excess of global glomerulosclerosis (median 49% of glomeruli, range 17-62%) when compared with the degree of interstitial fibrosis and tubular atrophy. Segmental sclerosis was less conspicuous (median 14%, range 6-31%). Vascular pathology predominantly affected arterioles.

## Discussion

We have previously reported that the clinical phenotype in Cypriot families with a hematuric nephropathy is more typical of a tubular disease until patients reach CKD stage 4 [8]. We have now confirmed this clinical phenotype in 20 such families with no pathogenic variants in *COL4A3*, *COL4A4* or other Mendelian glomerulopathy genes such as, *INF2* and *PAX2*. For convenience, we chose families with either the p.G545A or p.G999E variants of *COL4A4* but families without these variants behave identically.

In our experience, the earliest features of this Cypriot nephropathy are more consistent with a tubular disease as characterized by little or no proteinuria, and histology that shows widespread glomerular sclerosis, occasional segmental sclerosis but otherwise normal glomeruli with extensive tubular atrophy and interstitial fibrosis. These are the non-specific features of benign nephrosclerosis [10]. Unfortunately, electron microscopy was not available with the exception of one case in which the GBM showed pathological thinning in a patient with no pathogenic variant [8].

The weakness of our study is that we have not yet found a monogenetic explanation, although our hypothesis remains that the pathology is related to variants in *COL4A3*, *COL4A4* and *COL4A5*. It is unfortunate that we have not been able to do electron microscopy on our biopsies with one exception that supports our hypothesis. In this one case there was characteristic thinning of the GBM although no pathogenic variant was found [8]. Recently, Riella and his colleagues in Boston have reported that only one third of 49 patients reviewed with microscopic hematuria and thin glomerular basement membrane disease had a pathogenic variant of *COL4A3*, *COL4A4* and *COL4A5* [11]. This is entirely consistent with our experience of strong clinical evidence of an Alport nephropathy pathology but lack of genetic evidence - at least so far in the exons. Furthermore, this clinical presentation is very similar to that of the autosomal dominant *COL4A3* and *COL4A4* nephropathy described in Greek Cypriots, in whom under the age of 30, the only laboratory abnormality reported was microscopic hematuria and thin GBM disease and yet pathogenic variants were found in only 28% of families [5].

We suggest that hematuric nephropathy is part of an Alport spectrum nephropathy that now extends to a tubular phenotype. Recent experimental data has confirmed a role for tubules in the renal pathogenesis. Apart from the GBM, equally strong expression of collagen α(IV) chains is found along the distal renal tubule, and histology and primary tubular cell cultures of patients with Alport syndrome have demonstrated that in the tubular basement membrane the collagen IV molecule (α345) is largely produced by the distal tubule [12].

Tubular disease is common in Cyprus and in Greek Cypriot patients with tubular disease is usually related to variants in either protein kinase D1/2 (*PKD1/2*), mucin 1 (*MUC1*) or uromodulin (*UMOD*) [13]. We have also studied families with a tubular phenotype and only rarely have we found *UMOD* or *MUC1* mutations. Twelve patients that we investigated for hepatocyte nuclear factor 1-beta (HNF1B) because of a tubular phenotype with diabetes and cystic kidney disease produced no positive result [14].

We have observed that the presentation of Cypriot nephropathy resembles so-called hypertensive nephropathy. Hypertensive nephropathy is a controversial condition [15,16]. When the term hypertensive nephropathy is used it usually refers to a patient with hypertension and slowly progressive kidney disease, with minimal proteinuria and a biopsy that shows no evidence of glomerular disease apart from extensive glomerulosclerosis (benign nephrosclerosis) similar to our families [10]. Many nephrologists believe it does not exist as a definable kidney disease. Here we present evidence that there can be a genetic cause for this clinical phenotype.

We also propose that Cypriot nephropathy is likely to account for the great excess of kidney failure in the eastern Mediterranean in those over 65 years of age. Reviewing the annual Council of the European Renal Association and European Dialysis and Transplantation Association (ERA-EDTA) data on geographical variation in incidence of ESKD, Cyprus has an incidence of 3–4 times more common than in UK. But when the data is examined by age groups it can be seen that the Cyprus excess is predominantly owing to the much higher incidence after age 60, and in the age group 75+ the difference is 5-times compared with UK (Fig 2) [2].

**Figure 2.**
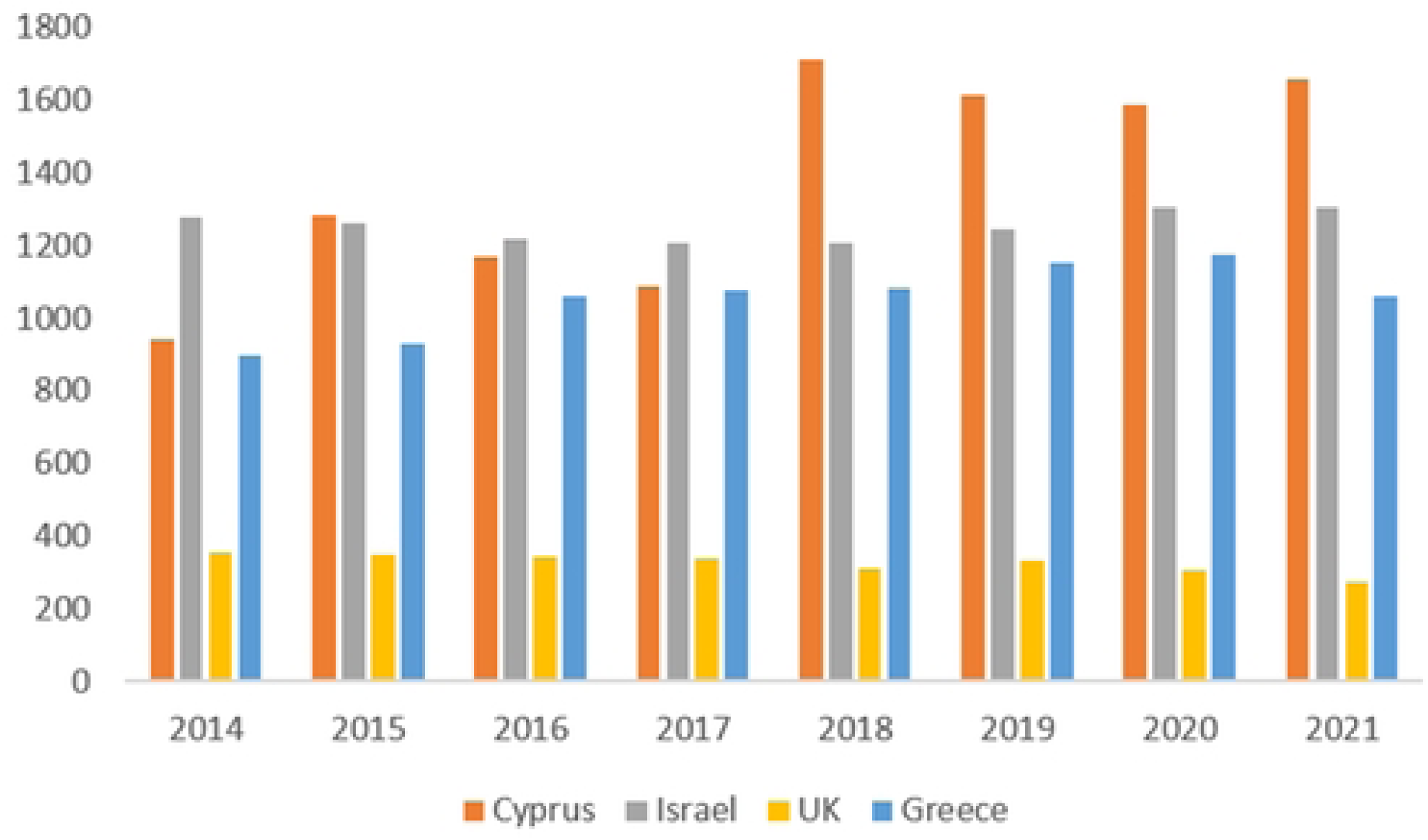
Incidence per million population (pmp), (age-related) by age 75+ unadjusted [2].

Our data suggests that Alport spectrum nephropathy extends to a tubular phenotype and recent experimental data has confirmed that this hypothesis is consistent with the renal pathogenesis [12]. But further studies will be needed to determine if the pathology relates to variants in Alport genes. More extensive genetic investigation is required to exclude deletions, insertions, copy number variation and intronic variants in these genes. We should consider that some other gene variant that is involved in GBM assembly may pay a role, such as the transcription factor LMX1B, which regulates the expression of collagens IV alpha 3- 5 network, or enzymes that are involved in collagen cross-linking such as peroxidasin (PXDN).

## Data Availability

The data that support the findings of this study are not publicly available as they contain information that could compromise the privacy of research participants but are available from the corresponding author upon reasonable request.

## Acknowledgements

We would like to thank the families included in the study.

## Author Contributions

Research idea and study design: GHN, DDO, CG; clinical care, data and sample collection: DDO, AB, SA, MY, SO, FO; laboratory studies: SO, FO, HHK, CG; data analysis/interpretation: FO, GHN, HHK, CG; statistical analysis: GHN, CG; manuscript writing: GHN, DPG, CG, HHK.

